# Contraceptive Outcomes of the Natural Cycles Birth Control App: A Study of Canadian Women

**DOI:** 10.1101/2024.09.13.24313618

**Authors:** Eleonora Benhar, Agathe van Lamsweerde, Kerry Krauss, Elina Berglund Scherwitzl, Raoul Scherwitzl

**Affiliations:** Natural Cycles Nordic AB, Sankt Eriksgatan 63 B, 112 34 Stockholm, Sweden

## Abstract

**Objective:** This study aimed to investigate the key demographics and evaluate the real-world contraceptive failure and continuation rates of the Natural Cycles app in a cohort of women from Canada.

**Methods:** This was a real-world, prospective cohort study. Demographics were assessed via in-app questionnaires. Contraceptive failure rates in typical and perfect use were calculated using the 13-cycle cumulative pregnancy probability (Kaplan-Meier survival analysis) and the one-year Pearl Index (PI). One-year continuation rates were estimated through survival analysis.

**Results:** The study included 8 ‘798 women who contributed an average of 9.2 months of data, amounting to a total of 7’ 063 woman-years of exposure. The average user was 27.3 years old, had a body mass index of 24.6, and reported being in a stable relationship. With typical use, the app demonstrated a 13-cycle cumulative pregnancy probability of 4.8 [95% CI: 4.3, 5.4] and a Pearl Index of 4.3 [95% CI: 3.9, 4.8]. Under perfect use, the contraceptive failure rate was 2.3 [95% CI: 0.7, 3.9] for life table analysis and 1.7 [95% CI: 0.5, 2.8] for the 1-year PI. The contraceptive method’s continuation rate after one year was 62.4%.

**Conclusions:** The data presented in this study offer valuable insights into the cohort of women using the Natural Cycles app in Canada and provide country-specific effectiveness estimates. The app’s contraceptive effectiveness aligns with previously published data on Natural Cycles.

## Introduction

Women in Canada are progressively favoring contraceptive methods with lower typical use effectiveness such as condoms and withdrawal, as shown by a study conducted by the Society of Obstetricians and Gynaecologists of Canada. The use of oral contraceptives has decreased by more than half over the last 10 years in women over 30, with side effects such as weight gain and headaches listed as the most common reasons for discontinuation. One quarter of women who stopped oral contraceptives for any reason turned to withdrawal or had unprotected sex. Another cause for concern is found in the fraction of women under 30 years of age lacking any contraceptive form growing significantly in the last decade [1]. Younger age, living in rural and remote territories, recent immigration, and lower socio-economic status all predicted higher dissatisfaction with current contraceptive options [2]. Accordingly, voices in the field are calling for lowering medical, financial, and educational barriers to contraception among Canadian women [3–5].

These findings emphasize the importance of expanding contraceptive options or facilitating access to meet the needs of this diverse population of 8 million women of reproductive age, particularly among those who are turning away from hormonal methods and may prefer non-hormonal and non-invasive options. Fertility awareness-based methods (FABM), which involve tracking physiological indicators such as basal body temperature or cervical mucus, are recently gaining traction as contraceptive alternatives [6]. The FDA-cleared digital birth control app Natural Cycles represents an innovative advancement in FABMs. It uses a smart algorithm to predict ovulation and fertility by analyzing user-entered temperature data and menstrual cycle patterns. Moreover, Natural Cycles addresses common barriers to FABM use by integrating with popular devices like the Apple Watch and Oura Ring, which reliably and seamlessly measure temperature during the night, and automatically adjusting for factors such as sleep disturbances or alcohol consumption. By offering a non-hormonal, science-backed, and evidence-based approach to contraception and fertility planning [7–9], Natural Cycles provides women with a modern and accessible solution. It is FDA-cleared and approved for use as a contraceptive in Europe, Canada, Australia, Singapore, and South Korea.

Reliable regional data on contraceptive effectiveness of this emerging class of digital methods is important for prospective users and medical professionals in Canada to make informed choices. This study provides a comprehensive evaluation of the effectiveness and user engagement with the Natural Cycles app throughout Canada, in a real-world setting considering a diverse range of user characteristics and behaviors.

## Material and methods

### The mobile application

Natural Cycles is a mobile application designed for contraception and fertility monitoring. By analyzing users’ input of basal body temperature and menstruation dates, it estimates the most likely upcoming ovulation date, fertile window, and subsequent menstruation dates. Optionally, users can perform urine luteinizing hormone (LH) test during the days approaching predicted ovulation and this information will be taken into account by the algorithm’s calculations. As a regulated medical device, the Natural Cycles app is cleared by the FDA in the United States and certified to be used as a contraceptive in Europe, Canada, Australia, Singapore, and South Korea.

Upon the users’ daily input, the application analyzes each individuals’ data with a statistical algorithm to predict the likelihood of fertility on any given day, and then indicates the users’ fertility status through a color-coded system. A day predicted as fertile is shown in “red”, and the user is instructed to “Use protection”. On a day predicted as unlikely to be fertile, shown as “green”, the user is told to be “Not fertile”. Previous studies demonstrated the estimation of ovulation by the algorithm to be highly precise [7]. In a global cohort of 22’ 785 women, the typical use Pearl Index was estimated to be at 6.9 [95% CI: 6.5, 7.2] [8]. Country-specific estimations were also obtained for users in the United States and in the United Kingdom [9,10]. The one year typical use Pearl Index in each country was found to be respectively 6.2 [95% CI: 5.5, 7.0] over 5’879 women and 6.1 [95% CI: 5.6, 6.6] over 12 ‘247 women.

The detection of ovulation by Natural Cycles relies on evaluating changes in basal body temperature over time, which implies obtaining stable daily readings of their temperature. During the period studied by this analysis, users could track their temperature using either a two-decimal basal thermometer or the Oura Ring, a wearable device that monitors physiological data. Users that measure their temperature manually with a thermometer are told to measure approximately at the same time each day (within a 4-hour window around their usual wake up time), as soon as they wake up (before any other physical activity such as sitting up, drinking water, snoozing, or getting out of bed) and only once per day. The recommendation for optimal app experience is to aim for adding at least five temperatures per week, or 70% of the days in other terms. Users are encouraged and supported with in-app notifications, reminders, and tips.

### Study design

This study is a real-world, prospective observational analysis of users of the Natural Cycles app who registered in Canada with the intent of preventing pregnancy. The analysis includes data from users who subscribed between January 1, 2020, and July 1, 2023. The dataset was finalized on July 1, 2024, ensuring all participants had the potential to contribute up to 13 cycles or one year of data.

We excluded users who logged fewer than 20 days of data, including recordings of menstruation, basal body temperatures, urine test results, or symptoms. Users who reported using long-acting contraceptives, such as copper intrauterine devices (IUDs), vasectomies, or tubal ligations, were also excluded. Women must be at least 18 years of age to make a purchase of a Natural Cycles subscription, so no minors were enrolled in this trial. Since Natural Cycles is incompatible with hormonal contraception, users of these methods could not register and were not included.

There were no exclusions based on cycle length, regularity, measurement frequency, or body mass index (BMI) for registering on the NC° Birth Control app or inclusion in this analysis.

User demographics were collected both during registration to the app and through additional in-app surveys during use.

### Data analysis

This analysis evaluates the typical use and perfect use contraceptive failure rates of the Natural Cycles app, along with the contraceptive continuation rate at one year.

Typical use includes all cycles and unintended pregnancies, including users who engaged in unprotected sex or relied on withdrawal on a “red day” (a day identified as fertile by the app). This measure reflects real-life usage of the contraceptive.

Perfect use is assessed in users who reported using either abstinence or barrier protection (e.g., condoms or diaphragms) on red days, as per the app’s Instructions For Use. Only cycles where the intercourse data logged in the app aligns with these methods are included in this evaluation.

This analysis reports the contraceptive effectiveness with both the Pearl Index and cycle-by-cycle survival analysis. More precisely the Kaplan-Meier estimate of the 13-cycle contraceptive failure rate is given [11].

The contraceptive continuation rate is defined as the probability that users will continue using NC° Birth Control after one year. This is calculated through per-day survival analysis, with the event surveyed being discontinuation, whether due to switching to other contraceptive methods or transitioning to planning a pregnancy instead with NC° Plan Pregnancy.

Data extraction and analysis were conducted using Kotlin and Python programming languages. Natural Cycles is compliant with GDPR standards, and researchers only had access to pseudonymized data [PRIVACY POLICY]. An ethics waiver was obtained from the Reading Independent Ethics Committee for the analysis of de-identified data.

### Determining pregnancy and discontinuation

Users who completed 13 cycles or one year on NC° Birth Control without an unintended pregnancy were right-censored.

Pregnancy was confirmed if users reported a positive pregnancy test, switched to NC° Follow Pregnancy mode, or confirmed an unintended pregnancy via follow-up email. If a participant stopped logging data late in the luteal phase with high sustained temperatures and did not respond to follow-ups, pregnancy was assumed. Users who discontinued earlier in their cycle were censored at the end of their last completed cycle. The same applied to those who switched to NC° Plan Pregnancy or reported an intended pregnancy.

### Patient Involvement

No patients were directly involved in this study as only anonymised or aggregated data were used. However, the effectiveness of this app as a method of contraception is an area of significant interest to current and prospective users and medical professionals in Canada.

Natural Cycles is compliant with GDPR standards: all sensitive data is anonymised and all personal data that can identify a user is deleted three years after an individual stops using the service. In addition, Natural Cycles has recently developed and released a full anonymisation functionality, such that an individual who perceives an increased risk can choose to use the app with full identity protection.

## Results

### Study Population

The study included 8 ’798 women who contributed an average of 9.2 cycles of data, totaling 7 ’063 woman-years of exposure. The cohort had a mean age of 27.3 years, with the majority of users below 35 years old (91.1%). The body mass index (BMI) ranged from 13.6 to 61.2, with a mean of 24.6, and 12.5% of respondents were classified as obese (BMI > 30.0). Most women (90.9%) did not have a medical condition at registration, but 1.9% indicated endometriosis, 2.9% thyroid disorders, 3.9% polycystic ovary syndrome, and 0.5% peri-menopausal symptoms.

The average cycle length in the dataset was 30.2 days long. Among this cohort of Canadian women, 27% met or exceeded the official recommendation of 70%, 19% measured between 50% to 70% of days and 54% added temperature for up to 50% of days.

When reported, most participants indicated being in a stable relationship (85.4%) and having a university degree or higher (73%) (Table 1). The majority of respondents reported being nulliparous (88.7%). The most common contraceptive methods used prior to adopting Natural Cycles were hormonal birth control (64.2%), barrier protection (12.9%) and withdrawal (8.4%) (Table 2). In this cohort of Canadian women, 8.3% did not have a contraception method before using Natural Cycles. Less commonly used methods included fertility awareness-based methods and a copper intrauterine device.

**Table 1:** Sociodemographic characteristics reported by study participants.

| Characteristic | Percentage of respondents (number) |
| --- | --- |
| **Age in years (mean ± std)** | *27.3 ± 5.1* |
| 18-24 | 31.2% (2746) |
| 25-29 | 39.7% (3487) |
| 30-34 | 20.2% (1774) |
| 35-39 | 6.5% (574) |
| $\geq 40$ | 2.4% (212) |
| **BMI (mean ± std)** | *24.6 ± 5.1* |
| <18 | 2.2% (83) |
| [18, 25[ | 62.4% (2367) |
| [25, 30[ | 22.9% (869) |
| >30 | 12.5% (472) |
| **Relationship status** |  |
| Long-term commitment or married | 58.9% (1'550) |
| In a relationship | 26.5% (696) |
| Complicated or other | 2.8% (74) |
| Single | 11.8% (310) |
| **Education level** |  |
| Elementary school | 0.3% (7) |
| High school degree | 11.0% (257) |
| Trade/technical/vocational training | 15.8% (369) |
| University degree | 70.4% (1649) |
| PhD, MD and/or JD | 2.5% (59) |
| **Medical condition** |  |
| None | 90.9% (7993) |
| PCOS | 3.9% (342) |
| Thyroid disorder | 2.9% (259) |
| Endometriosis | 1.9% (164) |
| Perimenopause | 0.5% (40) |

**Table 2:**
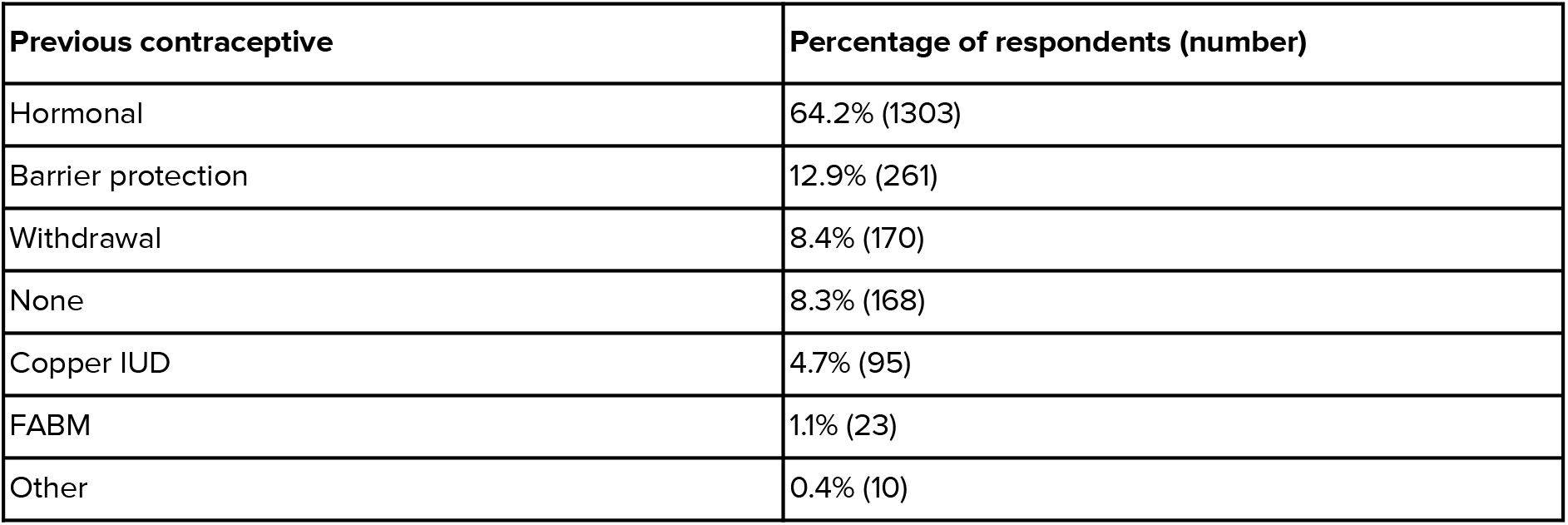
Contraceptive methods used prior to adopting Natural Cycles.

| Previous contraceptive | Percentage of respondents (number) |
| --- | --- |
| Hormonal | 64.2% (1303) |
| Barrier protection | 12.9% (261) |
| Withdrawal | 8.4% (170) |
| None | 8.3% (168) |
| Copper IUD | 4.7% (95) |
| FABM | 1.1% (23) |
| Other | 0.4% (10) |

When asked about their chosen method of protection on days flagged as fertile by the app, 52.5% of users reported using condoms, 27.7% relied on withdrawal, and 9.0% practiced abstinence (Table 3). However, only 16.6% of participants responded to this question. As a result, with 61.5% of respondents adhering to the instructions by using barrier protection or abstinence, 5 ‘514 cycles (7% of all cycles) from 1 ‘085 women (12% of all participants) were confirmed to be compliant with perfect use.

**Table 3:** Choice of birth control for “red” days (days indicated as fertile on the app, with the mention “Use protection”) reported by study participants, as answer to the question “*Which type of protection do you use on Red Days?*”.

| Type of protection used on red days | Percentage of respondents (number) |
| --- | --- |
| Condom | 52.5% (767) |
| Withdrawal | 27.7% (404) |
| A combination | 9.2% (135) |
| Abstinence | 9.0% (132) |
| None, I risk it | 1.2% (17) |
| Diaphragm or female condom | 0.3% (4) |
| Spermicide | 0.1% (2) |

### Contraceptive failure rates and continuation rates

At the end of the study, 306 (3%) women were determined pregnant, 2 ‘900 (33%) discontinued use of the app as birth control and 5’ 592 (64%) participants were still using Natural Cycles as birth control (Figure 1). Among those who discontinued, 6% did so to begin planning a pregnancy. Of the 306 women labeled as pregnant, 86% reported a positive pregnancy test in the app, 5% switched to the pregnancy monitoring mode in the app, 7% confirmed the pregnancy via follow-up email, and 3% were presumed pregnant by the app’s algorithm in the absence of user confirmation, based on late menstruation and consistently high temperatures. Among Natural Cycles users canceling the app’s subscription, the most frequent reasons given were “I’m trying to save money” (28%), “I no longer need Natural Cycles” (10%), and “It doesn’t work for me” (9%), with 18% deciding not to answer.

**Figure 1:**
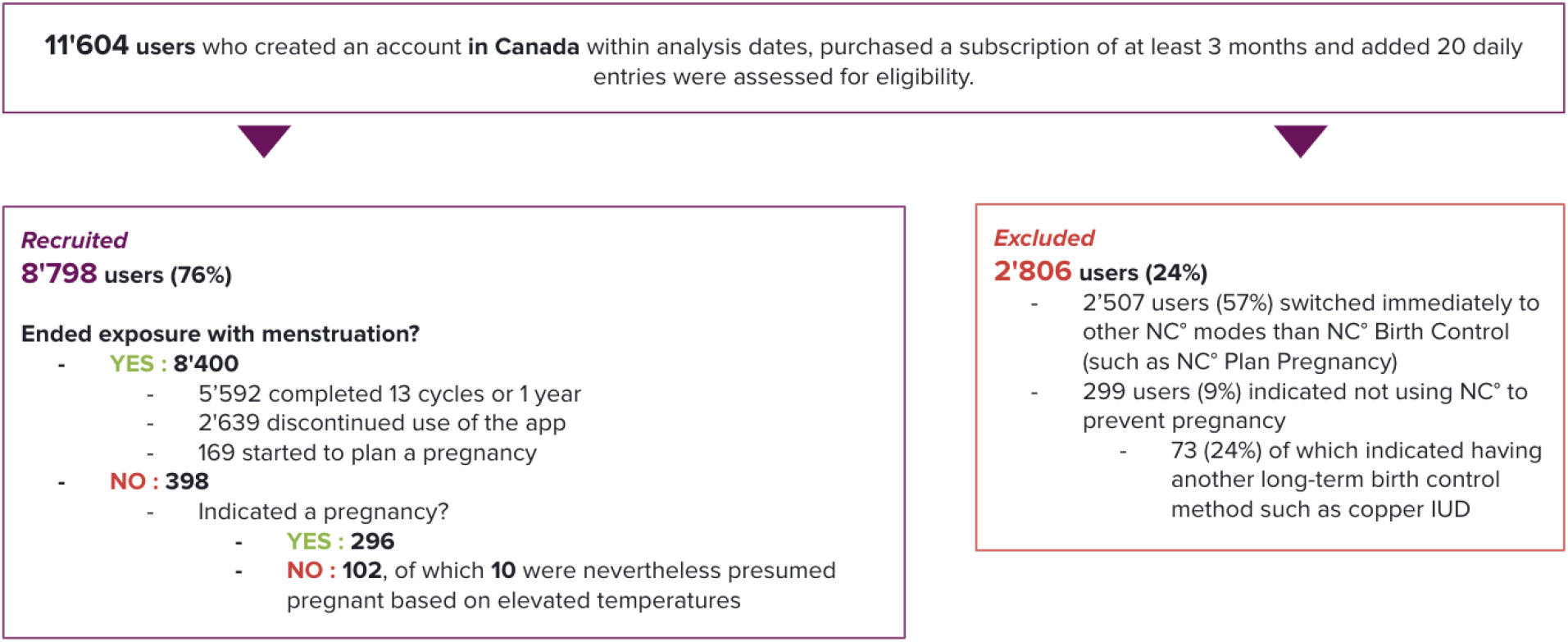
STROBE diagram of patients’ selection

With typical use, the app demonstrated a 13-cycle cumulative pregnancy probability of 4.8 [95% CI: 4.3, 5.4] and a PI of 4.3 [95% CI: 3.9, 4.8]. Under perfect use, the contraceptive failure rate was 2.3 [95% CI: 0.7, 3.9] for life table analysis and 1.7 [95% CI: 0.5, 2.8] for the 1-year PI.

The continuation rate for the method after one year was 62.4%.

## Discussion

The use of apps for fertility tracking and pregnancy prevention is becoming increasingly popular. Natural Cycles is the only app certified as a contraceptive in Canada, making research on its effectiveness within this specific cohort crucial for both users and healthcare providers when discussing contraceptive options.

With typical use, the app demonstrated a 13-cycle cumulative pregnancy probability of 4.81% [95% CI: 4.26%, 5.35%] and a PI of 4.33 [95% CI: 3.85, 4.82]. Under perfect use, the contraceptive failure rate was 2.25% [95% CI: 0.66%, 3.85%] for life table analysis and 1.68 [95% CI: 0.51, 2.84] for the 1-year PI. These results are comparable to, or slightly better than, previously published effectiveness rates [8–10].

The continuation rate for the method after one year was 62.4%. In comparison, a review of other fertility awareness-based methods (FABMs) reported 12-month continuation rates of 46% for the Standard Days Method and 52.7% for the TwoDay Method [6]. The contraceptive continuation rate for oral contraceptive pills can vary based on factors and was estimated to be between 46-55% [12,13].

### Strength and weaknesses of the study

Data were collected prospectively in a real-world setting from a large cohort of Canadian women. Pregnancy testing was conducted at home by the users themselves. To minimize loss to follow-up as some individuals discontinued the app, multiple push notifications and emails were sent to inquire about their pregnancy status, making it easy for users to report a possible pregnancy.

The analysis of real-world users reveals why a significant portion of socio-demographic data is missing, as answering these questions is entirely voluntary within the app. This issue also affects the question regarding the type of protection used on red days, which is essential for defining perfect use of the product. In this study, only 16.6% of participants responded to this question. As a result, although 53% of respondents reported using barrier protection and 9% practiced abstinence, many compliant users may not have been captured due to incomplete responses.

A key limitation of this analysis is the lack of reliable and consistent data on sexual intercourse. Notably, 81 pregnancies (26.5%) occurred in cycles where no intercourse information was reported by the user. Future efforts will focus on enhancing in-app tracking of sexual activity to address this gap and to allow for the calculation of effectiveness based solely on cycles where vaginal intercourse is confirmed.

## Conclusions

This study outlines the contraceptive outcomes and key demographics of a cohort of users from Canada. Digital fertility awareness-based methods are increasingly becoming a viable option for women of reproductive age, making it essential to generate country-specific scientific evidence on the effectiveness of these methods.

With typical use, the app showed a 13-cycle cumulative pregnancy probability of 4.8 [4.3, 5.4] and a Pearl Index of 4.3 [3.9, 4.8]. Under perfect use, the contraceptive failure rate was 2.3 [0.7, 3.9] based on Kaplan-Meier survival analysis and 1.7 [0.5, 2.8] for the 1-year PI. These effectiveness rates are in line or improving on previously published analyses of the Natural Cycles app in other countries [8–10].

## Data Availability

All data produced in the present study are available upon reasonable request to the authors.

## Author roles, conflicts of interests and funding

### Author contributions

AvL: Conceptualization, Methodology, Data Curation, Software, Formal analysis, Writing - Original Draft

EB: Conceptualization, Methodology, Supervision, Writing - Original Draft

KK: Conceptualization, Writing-Reviewing and Editing

EBS: Conceptualization, Software, Writing-Reviewing and Editing

RS: Conceptualization, Writing-Reviewing and Editing

### Funding

Natural Cycles provided access to anonymised data collected on its digital fertility application as well as salaries for AvL, EB, KK, EBS and RS as employees.

### Data availability

The datasets generated and/or analyzed during this study are not publicly available due to the proprietary nature of the algorithm and complexity of outputs but are available from the corresponding author on reasonable request.

## Abbreviations

(BMI): Body mass index
(FABM): Fertility awareness based methods
(FDA): US Food and Drug Administration

